# White Matter Microstructure in Habit and Reward Circuits in Anorexia Nervosa: Insights from a Neurite Orientation Dispersion and Density Imaging Study

**DOI:** 10.1101/2022.07.20.22277868

**Authors:** Stuart B. Murray, Ryan P. Cabeen, Kay Jann, Reza Tadayonnejad, Michael Strober, Jamie D. Feusner

## Abstract

**Background:** Behavioural features of anorexia nervosa (AN) suggest abnormalities in reward and habit. Neuroimaging evidence suggests morphometric and functional perturbations within these circuits, although fewer studies have assessed white matter characteristics in AN, and no studies to date have assessed white matter microstructure in AN.

**Methods:** In this brain imaging study, 29 female adolescents with partially or fully weight-restored AN and 27 healthy controls, all between 10-19 years, underwent whole-brain multi-shell diffusion tensor imaging. Utilizing neurite orientation dispersion and density imaging methods, we investigated group differences in white matter neurite density, orientation dispersion, and myelin density in tracts between prominent nodes of the reward circuit (ventral tegmental area (VTA) to nucleus accumbens (NAcc)) and the habit circuit (sensory motor area (SMA) to putamen).

**Results:** Findings revealed reduced neurite (F=5.20, p=0.027) and myelin density (F=5.39, p=0.025) in the left VTA-NAcc tract, and reduced orientation dispersion in the left (F=7.00, p=0.011) and right (F=6.77, p=0.012) VTA-NAcc tract. There were no significant group differences in the SMA-putamen tract. Significant relationships, after corrections, were not evident between tract microstructure and reward responsiveness, compulsive behaviours, illness duration, or BMI.

**Conclusions:** Adolescents with AN exhibit less dense, undermyelinated, and less dispersed white matter tracts connecting prominent reward system nodes, which may signify underutilization of this part of the reward circuit. These results provide a detailed examination of white matter microstructure in tracts underlying instrumental behavioral phenotypes contributing to illness in AN.

---

Anorexia nervosa (AN) is a pernicious psychiatric disorder which is principally characterized by rigid dietary restriction and a fear of weight gain, often despite a dangerously low body weight (1). Alongside a homogenous symptom profile and illness course, AN is additionally characterized by a stereotypic age of onset during early adolescence, a female gender predominance, and a mortality rate that exceeds that of other serious psychiatric disorders (1). Importantly, the pathophysiology of AN remains unknown, and neurobiological models that adequately explain the symptom domains have remained elusive. Explicating the neurobiological mechanisms associated with the key phenomenological features of AN is critical in developing more effective treatments (2, 3).

A confluence of evidence has revealed abnormalities in what may be two important neural and behavioural phenotypes in many with AN: reward learning and habitual decision making. Behaviorally, those with AN typically report lower reward sensitivity (4), greater anhedonia (5), lower novelty seeking (6), and demonstrate an elevated capacity to delay reward seeking (7, 8), which extends to both food and broader hedonic cues. As well, those with AN also demonstrate greater proclivities towards habit-based decision making (e.g., rigid and continued engagement in behaviours such as restricted food selection and exercise routines) (9). Collectively, these abnormalities are thought to underpin (i) the characteristic capacity to restrict palatable foods, which are typically experienced as hedonic and rewarding to others, and (ii) the persistence of symptomatic behaviours, often despite intensive treatment and the disconfirmation of disorder-specific fears (i.e., exponential weight gain). With respect to habit-based decision making in AN, evidence has illustrated that the degree of habit strength is associated with a longer duration of illness (10) and is predictive of symptom severity (11).

Two neurobiological models have been advanced to account for these abnormalities – (i) one relating to reward (12-14), postulating widespread dysfunction in frontostriatal and mesolimbic dopaminergic pathways (15-19) that causes pervasive under-responding to cues typically deemed hedonic, and (ii) one relating to habit learning (3, 9), postulating dysfunction in dorsal frontostriatal networks, and the development of rigid and ritualized eating behaviors. Functional studies have found abnormalities in reward and habit-related regions in those with AN, although the directionality of these findings has been mixed to date (13, 20-23). Structural studies assessing white matter tracts have also revealed widespread abnormalities in tract integrity (24), although studies to date have focused largely on major white matter pathways, as opposed to hypothesis-driven seed-to-seed analyses. Our group’s recent seed-to-seed tractography analyses of white matter tracts between sensory motor area (SMA) to putamen, a circuit known to be implicated in habitual decision-making (25), found that the volume of tracts in this circuit were associated with the severity of ritualized behaviors (25). A separate study revealed delayed response time in adolescents with AN when anticipating potential rewards (26). Also in that study, probabilistic tractography illustrated reduced integrity in white matter tracts connecting the ventral tegmental area (VTA) and the nucleus accumbens (NAcc) – a major mesolimbic dopaminergic projections pathway implicated in reward. As well, there was reduced VTA-NAc functional connectivity in those with AN. An important next step in this line of inquiry lies in more completely understanding abnormalities in these white matter tracts on the microstructural level, and their relationship to AN symptomatology.

With the advent of more sophisticated analytical methods of discerning white matter microstructure, this study represents the first known application in AN of white matter neurite orientation and dispersion density imaging (NODDI) (27), focusing in particular on tracts relating to reward and habit. White matter microstructure has rarely been assessed in psychiatric populations, owing to its reliance on scarcely available postmortem tissue samples, although the recent development of NODDI offers a non-invasive and in-vivo method of assessing neurite morphometry (27, 28). Indeed, the quantification of neurite morphology, which collectively refers to dendrites and axons, offers important insights into the structural basis of brain function, development, and aging (27, 29, 30). With this novel method, abnormal neurite morphology has been illustrated in serious neurological and psychiatric disorders, including multiple sclerosis (31), Alzheimer’s Disease (32), and psychosis (33). The assessment of white matter microstructure is of particular relevance to AN, owing to the unique relationship between dietary fats – which is pervasively retracted in AN, brain lipids, and the development of myelinated white matter tracts.

Building on our group’s previous findings illustrating reduced white matter tract integrity in the (i) SMA to putamen and (ii) VTA to NAcc tracts (25, 26), we undertook NODDI analyses of these two tracts to interrogate both neurite density and neurite orientation. Moreover, we further leveraged a novel analytical model of absolute tissue density from NODDI (ABTIN) (34) to infer absolute myelin density. This mathematically derived model has demonstrated good concordance in inferences of absolute myelin density with that seen on electron microscopy histology (34). Per our preregistered hypotheses (https://osf.io/8kh26/), we predicted that those with AN would demonstrate (1) abnormal white matter neurite density and myelination in tracts connecting the SMA to putamen (i.e., habit circuit) and that abnormalities would be correlated with symptom-related compulsive behaviors. Moreover, we hypothesized that AN would be characterized by abnormal white matter neurite density, orientation dispersion, and myelination in tracts connecting the VTA to NAcc (i.e., reward circuit) and that myelination would be correlated with trait markers of reward responsiveness. Lastly, we hypothesized that myelination in both tracts would be associated with duration of illness.

## Method

### Participants and clinical evaluations

All participants were female adolescents, aged between 10-19 years of age who had a current DSM-5 diagnosis of AN, or were healthy controls with no lifetime criteria for any eating disorder (Table 1). AN participants were recruited from the inpatient Eating Disorders Unit at UCLA and residential/partial hospital treatment centers in Southern California and were either partially or fully weight-restored. Control participants were recruited through local advertisements. No history of head trauma was noted among any participants. Nineteen participants with AN were taking psychotropic medication, at a stable dose. This study was approved by the UCLA Institutional Review Board, and all participants provided assent, and informed consent was obtained from parents or legal guardians.

**Table 1:** The demographic and behavioral characteristics of the sample.

|  | **AN (n=29)** | **Control (n=27)** | **Statistic** | **P value** |
| --- | --- | --- | --- | --- |
| Age (years) | 14.8 ± 1.7 | 16.4 ± 1.8 | t=-3.36 | 0.001 |
| Education (years) | 8.8 ± 2.0 | 10.4 ± 1.9 | t=-3.20 | 0.002 |
| Pubertal status<sup>a</sup> | 14.5 ± 4.2 | 18.0 ± 1.5 | t=-4.0 | <.001 |
| BMI | 18.4 ± 1.4 | 22.6 ± 2.5 | t=7.61 | <.001 |
| BMI Percentile | 28.6 ± 17.4 | 63.5 ± 20.7 | t=-6.80 | <.001 |
| Duration of illness (months) | 19.9 ± 17 | NA |  |  |
| Psychotropic Medication (Y / N) | 19 / 10 | 0 |  |  |
| EDE – Total Score | 3.4 ± 1.5 | 0.4 ± 0.5 | t=10.23 | <.001 |
| EDE – Restraint | 2.8 ± 1.8 | 0.5 ± 0.7 | t=36.67 | <.001 |
| EDE – Restraint (6-month Follow-up)<sup>b</sup> | 2.3 ± 1.6 | NA |  |  |
| YBC – Total Score | 26.1 ± 9.1 | 1.3 ± 3.0 | t=13.86 | <.001 |
| YBC - Compulsions | 10.4 ± 4.2 | 0.3 ± 1.2 | t=12.45 | <.001 |
| YBC – Compulsions (6-month Follow-up) | 7.4 ± 6.3 | NA |  |  |
| BAS – Reward Responsiveness<sup>c</sup> | 16.5 ± 1.9 | 17.2 ± 2.1 | t=-1.21 | 0.232 |

<sup>a</sup>Missing data for 2 AN and 3 Controls<sup>b</sup>Missing data for 5 AN<sup>c</sup>Missing data for 2 AN and 1 Control

### Assessments

All participants underwent a structured clinical assessment (the Mini-International Neuropsychiatric Interview (MINI KID 7.0.2) (35) and the Eating Disorders Examination (36)) with a licensed psychiatrist or clinical psychologist specialized in the treatment of eating disorders. The Pubertal Development Scale was used to assess pubertal developmental stage (37). The Behavioral Inhibition System/Behavioral Activation System (BIS/BAS) (38) was administered, with the BAS reward responsiveness subscale used in analyses. The Yale-Brown-Cornell Eating Disorder Scale (YBC-EDS) (39) was administered to quantify eating disorder related obsessions and compulsive behaviours, with the compulsive subscale used in analyses.

### Diffusion Tensor Imaging Data Acquisition

Whole-brain data acquisition was undertaken with a Siemens PRISMA scanner using a 64-channel coil. An MPRAGE sequence was used to acquire T1-weighted structural data (TR = 2.3s, TE = 2.99 ms, and voxel dimensions .08 × .08 × .08 mm), and diffusion weighted imaging data were acquired using single-shot spin-echo echo-planar imaging (field of view=240mm; voxel size=1.5×1.5×1.5, with 0.75 mm gap; TR/TE=3,222/89ms and flip angle 89). We obtained 92 slices contiguous axial slices aligned to the AC-PC line along 99 gradient-sensitizing directions with b=3,000s/mm^2^, b=1500s/mm^2^, and seven minimally diffusion-weighted scans.

### Diffusion-Weighted Image Processing

Standard DWI processing was performed using a combination of the FSL Diffusion Toolbox (40), ANTs (https://stnava.github.io/ANTs/) (41), and the Quantitative Imaging Toolkit (QIT) (http://cabeen.io/qitwiki; 42). We first applied a non-local means filter to the DWI data using the VolumeFilterNLM module in QIT (43), and then FSL-eddy was used to correct for eddy current-induced distortions (44). An automated quality control step was performed to quantify the DWI data quality using FSL-eddy_quad (45) at a subject level. Brain segmentation was performed using FSL BET (46), and diffusion tensor imaging (DTI) parameters were estimated using weighted-linear least squares estimation (47) implemented in the VolumeTensorFit module of QIT, which provided a fractional anisotropy (FA) map. NODDI parameters were estimated using the spherical mean technique using the VolumeNoddiFit module in QIT (48), and these parameters were further processed to obtain ABTIN parameters using the VolumeNoddiAbtin module in QIT (34); these included estimates of myelin density, cerebrospinal fluid, fiber, and cellular densities. We computed a diffeomorphic warp field between each subject and the IIT template (49) based on FA and and ANTs SyN registration algorithm (41, 50), and we retained the warp field for later steps.

### Seed Selection

We performed a region-of-interest (ROI) analysis to obtain microstructure maps of the SMA-Put and VTA-NAcc pathways. For this, we defined these pathways in the IIT template using population averaged tractography. We defined the VTA region using the Caltech Reinforcement Atlas (51), and we manually drew an SMA ROI on a population averaged cortical surface. We used average putamen and nucleus accumbens ROIs obtained using Freesurfer (52). We performed deterministic streamline tractography using a population average multi-fiber diffusion template (53) to obtain connections between the putamen and SMA and between the nucleus accumbens and VTA. Tractography parameters included 20 seeds per voxel within each pair of ROIs, a maximum turning angle of 65 degrees, a minimum fiber volume fraction of 0.05, and strict termination of streamlines inside endpoint ROIs. We then computed a tract density map for each pathway, and we applied the warp field to deform the tract density map to each subject’s native space. We then computed a weighted average of each microstructure parameter, with weighting by the voxelwise tract density value.

### Data Analysis

Statistical analyses were performed using the MATLAB Statistics and Machine Learning Toolbox. We conducted 2-way ANCOVA for NAcc-VTA and Put-SMA fiber bundles, respectively, which controlled for age and pubertal status. Significance threshold was set at p<0.05 with Benjamini-Hochberg correction for multiple comparisons. Associations between behavioral assessments and microstructural tract properties were estimated for both groups separately by means of partial Spearman rank correlations, correcting for age and pubertal status. Significance was set at p<0.05. All hypotheses and analytical approaches were preregistered and are openly available at https://osf.io/8kh26/.

## Results

### Demographic and Behavioral Data

We obtained usable data from 29 participants with AN and 27 healthy controls. Significant between-group differences emerged for age and pubertal status (Table 1). Participants with AN also had lower age-adjusted BMI percentile (*p*<.001), greater EDE scores (*p*<.001) and greater YBC scores (*p*<.001). No significant differences in reward responsiveness were noted (*p*=.232).

### Brain Imaging Results

#### Microstructural differences within tracts

ANCOVA models controlling for age and pubertal status revealed significantly reduced NDI (F=5.20, p=0.027), ODI (F=7.00, p=0.011) and myelin density (F=5.39, p=0.025) in left VTA-NAcc tract in AN, and reduced ODI (F=6.77, p=0.012) in right VTA-NAcc tract in those with AN. No differences emerged in the SMA-Put tract emerged in NDI (left F=0.45, p=0.508; right F=0.29, p=0.594), ODI (left F=1.10, p=0.300; right F=3.29, p=0.076) or myelin density (left F=0.54, p=0.466; right F=0.34, p=0.595).

#### Association with Clinical Variables

The hypothesis-based correlation analysis with clinical variables in participants with AN demonstrated no significant associations between myelin in VTA-NAcc and reward responsiveness (left r=0.20, p=0.337; right r=0.23, p=0.272) or illness duration (left r=0.11, p=0.608; right r=0.28, p=0.170). Furthermore, there was no significant relationship between myelin in SMA-Put and eating disorder-related compulsions (left r=-0.19, p=0.362; left r=-018, p=0.389) or illness duration (left r=0.14, p=0.518; right r=0.10, p=0.636). Exploratory analyses beyond the hypotheses demonstrated a positive correlation between ODI in left VTA-NAcc and BAS reward responsiveness (r=0.43, p=0.033, uncorrected). Finally, in an exploratory analysis we found no significant relationships between any microstructural index and BMI-percentiles.

## Discussion

This represents the first study to estimate neurite density, orientation dispersion, and myelination of white matter tracts in AN. Using NODDI and ABTIN to probe white matter microstructure, we tested hypotheses relating to two neural systems that are relevant to prominent behavioral phenotypes in AN: reward processing and habitual decision-making. As predicted, in the reward circuit there was evidence for abnormal neurite density, orientation dispersion, and myelination in the VTA-NAcc tract in AN. However, and contrary to hypotheses, no group differences emerged in neurite density, orientation dispersion, and myelination in the SMA-put tract. We also did not confirm our hypotheses of associations between white matter microstructural indices and the clinical variables of eating disorder-related compulsive/ritualistic behaviors, duration of illness, or subjective reward responsiveness.

### Framework for understanding aberrant white matter microstructure in AN

The following provides context for what these results may signify about the white matter of adolescents with AN, on the cellular and molecular level, given the metabolic and developmental milieu of the illness. White matter is susceptible to malnutrition-related alterations that occur in the starvation state. While little is known about the effects of starvation on neurite density or orientation dispersion, several studies have explored the effects on myelination. Of the white matter microstructural elements, starvation and malnutrition may have the greatest effects on myelination (54), which informed our predictions for abnormalities in the AN group. One previous study that investigated white matter myelin content in underweight adolescent girls with AN using T1 relaxometry (55) found evidence for reduced myelin in the cingulate, corticospinal tract, inferior fronto-occipital fasciculus, and the arcuate fasciculus. Effects of undernutrition on myelination have also been demonstrated experimentally in rats. While most have examined the early postnatal period, one study used rats of an approximately equivalent developmental period as human early adolescence; myelin content was reduced as a result of both general undernutrition and protein malnutrition (56). Specifically, protein malnutrition resulted in both a greater reduction of myelin content and a reduction in 2’,3’-cyclic nucleotide 3’ phosphohydrolase (CNPase) and proteolipid protein, which was attributed to delays in maturation. In that study, decreases in myelin content were only partially restored by nutrition rehabilitation. Other studies have found persistent myelin phospholipid composition abnormalities resulting from malnutrition during developmental periods in rodents (albeit slightly younger), even after nutritional rehabilitation (57, 58). These animal findings are relevant to the population in the current study – adolescents with AN – since they are undergoing a similar stage of neurodevelopment and had recently undergone re-alimentation. We surmise that any perturbations of myelin from malnutrition may persist in this AN cohort; this could be due to the fact that they had only relatively recently achieved full or partial weight restoration, and/or that myelination-related brain maturation (occurring through adolescences and into the 20s (59)) may have been affected. Whether this recovers fully would need to be explored in future cohorts that are followed with longitudinal scans.

Notwithstanding this evidence for effects of malnutrition on myelination, another explanation for observed abnormalities in myelination, as well as neurite density and orientation dispersion, are that they may have pre-dated the starvation state. If so, such abnormalities could have been predisposing factors underlying the behavioural phenotypes of aberrant reward and habit formation that might contribute to the development of AN.

An additional possibility is that behaviours or other symptoms of the illness itself may have secondarily resulted in white matter microstructural abnormalities. The lack of significant associations between illness duration and myelination density argues against this possibility. Although, it cannot be ruled out completely due to most participants having undergone some form of treatment during the course of their illness prior to when data was collected, which confounds the effects of the illness itself with its associated treatments.

### White matter microstructural findings in reward circuit tracts

In the VTA-NAcc there was evidence for less dense, under-myelinated, and less dispersed tracts in AN than controls. As outlined, lower myelin density might signify an effect of starvation that incompletely resolved after re-alimentation, could represent a more permanent effect of malnutrition, could pre-date the starvation state as might occur with abnormal neurodevelopment, or could be a cause or effect of underutilization of this reward circuit pathway. Effects of starvation, as described above, could result in reduced myelin thickness and subsequent lower myelin density. Similarly, if starvation reduces neurite density, it could also secondarily result in reduced observed myelin density. How myelination density relates to reward-related subjective experiences is unclear, however, since significant (corrected) associations in the VTA-NAcc tracts with BAS reward responsiveness were not observed.

One speculation is that the set of findings in the AN group of less dense, under-myelinated, and less dispersed VTA-NAcc tracts might signify underutilization of this part of the reward circuit. This is supported by a previous analysis in the same sample of AN participants who engaged in a reward fMRI task, in which we observed reduced functional connectivity in VTA-NAcc and delayed response times for reward motivation – evidence for lower activity in this circuit (26). Further, in the current study, those with lower orientation dispersion in the VTA-NAcc – “straighter” fiber tracts – had lower BAS reward responsiveness. With the strong caveat that this was an exploratory finding that did not survive corrections for multiple comparisons, whether lower reward responsiveness could be a cause or effect of lower neural activity in this circuit remains to be determined. Notwithstanding, clinical studies assessing anhedonia in AN illustrate reliable elevations in the acute illness state, which are also evident in the long-term recovered state (60) suggesting that anhedonia (and perhaps the related phenomenon of diminished reward responsiveness) might persist independently of nutritional factors. In sum, while our specific hypotheses about correlations between myelination in the VTA-NAcc and reward responsiveness were not confirmed, potential behavioural and clinical relevance of less dense, under-myelinated, and less dispersed VTA-NAcc tracts in AN is underscored by related findings in these tracts in the same sample of reduced functional connectivity and delayed responses during reward motivation, as well as the exploratory finding in the current study of associations between straighter fiber tracts and lower reward responsiveness.

With regard to the habit tract, we found no significant differences in myelin density, neurite density, or neurite orientation in those with AN. In a previous examination of SMA-to-putamen tracts in a partially overlapping cohort of adolescents with AN and controls, we similarly found no significant differences in FA (nor in white matter volume or number of streamlines) (25). The present study also did not find significant differences, despite including a larger sample and examining multiple indices of WM microstructure rather than gross volume and tract number (which should afford greater sensitivity to detect white matter abnormalities associated with specific clinical conditions (55)). Cumulatively, these studies suggest that AN may not characterized by abnormalities in the white matter tracts between the SMA and putamen nodes of the habit circuit, at least in the weight-restored state.

Notwithstanding these findings, several limitations are noteworthy. First, the study was relatively modest in sample size. A larger sample size would optimize the capacity to reliably detect meaningful differences of a smaller effect size, in addition to being able to analyze subsets of the medicated (n=19) and unmedicated (n=10) participants. Secondly, this study focused on adolescence, which is the most common period of onset for AN (61). However, how these findings relate to adults with AN, who may have experienced longer periods of starvation, is unclear. This is particularly important to explore among adults of variable illness duration, given the plausible effects of starvation and malnutrition upon white matter characteristics (54). Moreover, despite our limited yet hypothesis-driven focus on habit and reward circuits, it is important to acknowledge the mounting evidence for abnormalities in circuits modulating fear (62), interoception (63), emotional regulation (64) and cognitive control (65) in AN, and additional inquiry ought to examine microstructural properties of these circuits.

In conclusion, these findings present novel insights about white matter microstructural abnormalities in a reward circuit in adolescents with AN: the tracts are less dense, undermyelinated, and less dispersed. Such abnormalities might be linked to characteristic clinical perturbations of reward experiences and previously described neural aberrancies in reward system functioning. On the contrary, abnormal white matter microstructure was not evident in habit circuit tracts. The results provide a detailed examination of white matter microstructure in tracts underlying instrumental behavioral phenotypes contributing to illness in AN. Longitudinal studies will be important to understand how these white matter characteristics contribute to the pathogenesis of AN.

**Figure 1:**
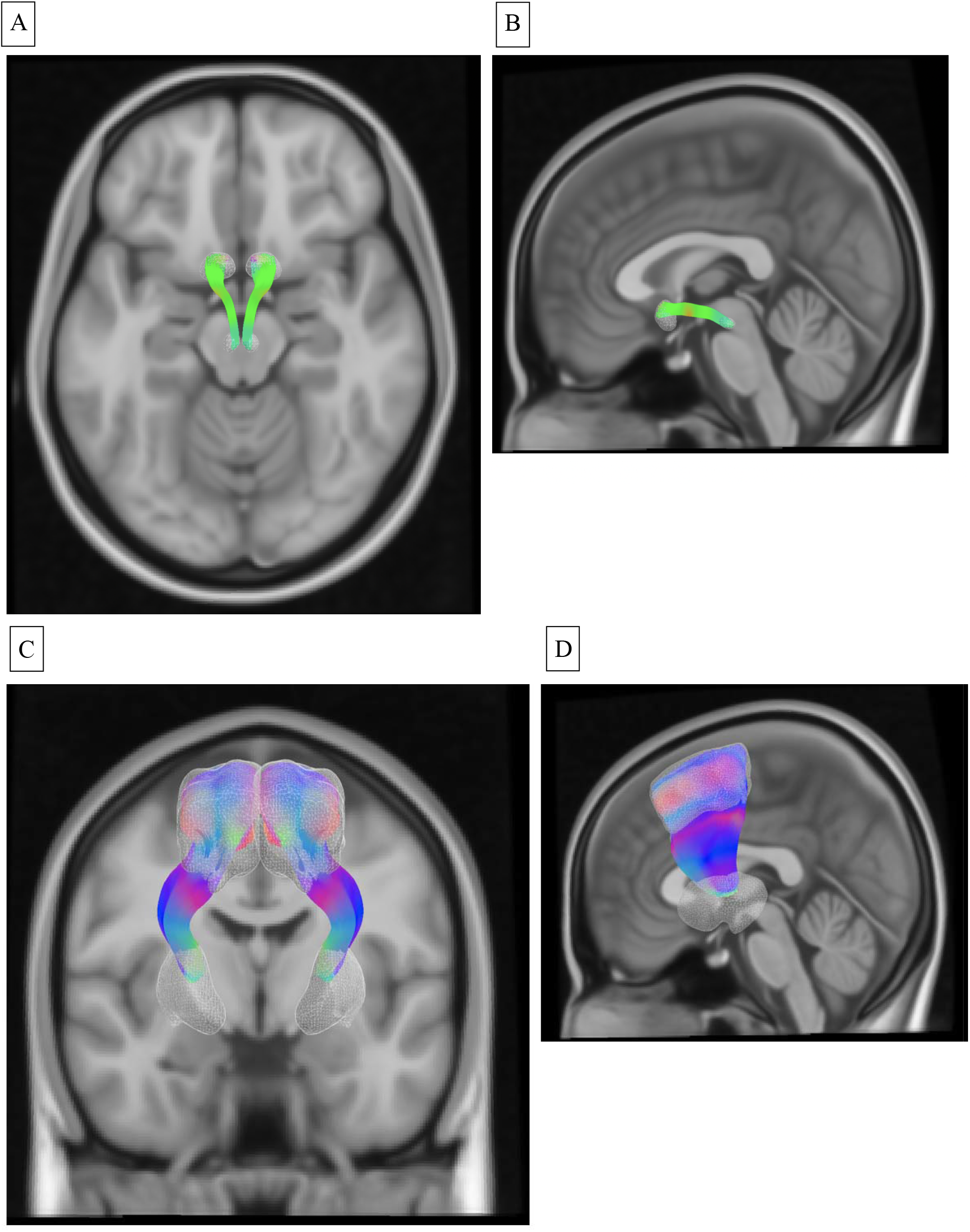
An illustration of the white matter tracts assessed, including the VTA-to-NAcc tract, shown from the axial (A) and sagittal (B) plane, and the SMA-to-putamen tract, shown from the coronal (C) and mid-sagittal (D) plane.

**Figure 2:**
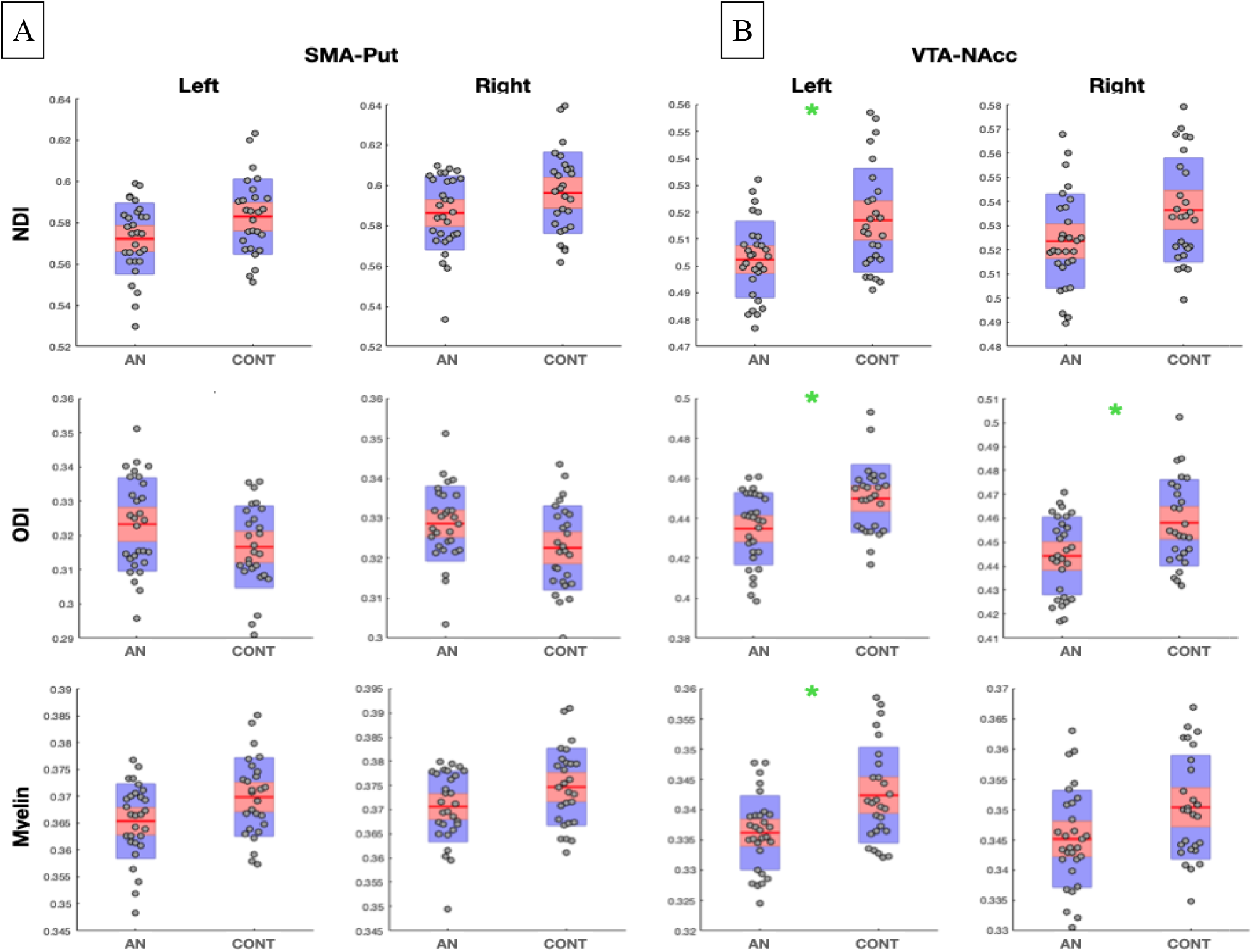
A visual illustration of group differences in (i) neurite density (NDI), (ii) neurite A B orientation dispersion (ODI), and (iii) myelin density among adolescents with AN (N=29) and controls (N=27) in the (A) SMA-to-putamen tract and (B) the VTA-to-NAcc tract.

## Data Availability

the data are available upon reasonable request to the Corresponding Author.

## Acknowledgments

The authors would like to thank the research participants for their interest and their generosity of time in participating in the study.

